# Titration regimens mitigate mocravimod-induced negative chronotropic effect while preserving the pharmacokinetic and pharmacodynamic properties

**DOI:** 10.64898/2026.07.07.26357458

**Authors:** Dymphy Huntjens, Dirk Klingbiel, Jens Hasskarl

## Abstract

**Background:** Sphingosine 1-phosphate receptor (S1PR) modulators can cause transient, dose-related negative chronotropic effects. Mocravimod is an oral S1PR modulator that is developed as a maintenance therapy in allogenic haematopoietic cell transplantation (allo-HCT). This phase I study evaluated whether two dose-titration regimens attenuate early bradycardia when initiating mocravimod while preserving pharmacokinetic (PK) and pharmacodynamic (PD) activity.

**Patients and methods:** In this randomized, double-blind, placebo-controlled, parallel-group study, healthy adults received once-daily oral mocravimod using either dose titration (DT) regimen DT1 (0.3–2.0 mg with 4-day stepwise escalation) or regimen DT2 (0.5 mg to Day 14, 1.2 mg Days 15–18, then 2 mg), a fixed 2 mg regimen, or placebo for 21 days. The primary endpoint was the number of bradycardia episodes on treatment initiation and dose-escalation days derived from 24-hour Holter monitoring; PK of mocravimod and mocravimod-phosphate (whole blood) and PD effects (absolute lymphocyte count [ALC]) were assessed.

**Results:** Fifty-six participants were randomized and 53 completed the study. Both titration regimens resulted in fewer bradycardia episodes than fixed initiation at 2 mg during the first week of treatment. Differences between titration and fixed dosing were no longer evident after Day 9, consistent with tolerance development. PK profiles were consistent with prior phase I data. By Day 21, DT1 achieved exposures close to the fixed 2 mg regimen, whereas DT2 yielded lower exposures, reflecting slower escalation. Peripheral lymphopenia developed in all active treatment groups and was comparable between regimens by Day 21, returning toward baseline by study end. Safety was similar between titration regimens and placebo, with similar distribution and incidence of adverse events. No serious adverse events occurred.

**Conclusion:** Two practical titration regimens mitigated the early negative chronotropic effect observed with fixed-dose initiation of mocravimod at 2 mg once daily. Importantly, titration preserved the expected PK and PD profile, supporting dose escalation as an effective initiation strategy to improve early cardiac tolerability.

## Introduction

Mocravimod is a novel, synthetic, second-generation oral sphingosine 1-phosphate receptor (S1PR) modulator. It is a pro-drug which is phosphorylated *in vivo* to its active metabolite, mocravimod-phosphate. Mocravimod-phosphate targets four out of the five S1PR isoforms with high agonistic potency on human S1PR1 and S1PR4 and only partial agonism on S1PR3 and S1PR5 [1].

S1PR modulators sequester lymphocytes into secondary lymphoid organs while attenuating their trafficking to the periphery, leading to peripheral lymphopenia as their primary pharmacodynamic effect [2]. In pathological states, it reduces lymphocyte trafficking to pathologically inflamed tissue, such as the central nervous system in multiple sclerosis or the gut in inflammatory bowel diseases. In patients who receive allogenic haematopoietic cell transplantation (allo-HCT), this mechanism helps to decouple the wanted graft-versus-leukaemia (GvL) effect from the unwanted graft-versus-host disease (GvHD) [2–4].

Early-phase studies have shown that mocravimod, consistent with other S1P receptor modulators, may cause an initiation-related reduction in heart rate [5, 6]. This effect is considered to result from initial S1PR1 signalling on cardiac myocytes, with activation of G protein-coupled inwardly rectifying potassium channels before receptor internalization and functional desensitization [7].

The single dose study CKRP203A2101 investigating doses ranging from 0.01 to 40 mg or placebo indicated that single oral doses of mocravimod up to 40 mg were safe and well tolerated in healthy adults, with predictable pharmacokinetic (PK) and expected pharmacodynamic (PD) effects [5].

In the multiple ascending dose study CKRP203A2102, once-daily mocravimod doses of 0.3, 0.6, 1.2, 2.0, and 3.0 mg were evaluated in healthy participants [6]. An initial, dose-related heart rate reduction was observed, with attenuation of this effect during continued therapy. At dose levels of 1.2–3.0 mg, mean heart rate reductions of approximately 10 beats per minute versus placebo were recorded during the first week of dosing, whereas smaller reductions were seen at 0.3 and 0.6 mg. In addition to bradycardia, asymptomatic but likely treatment-associated second-degree atrioventricular (AV) block Type I (Wenckebach) events were reported after dosing on Day 1 at the 2.0 and 3.0 mg dose levels. No Wenckebach events were recorded on Day 3 or later, despite a persisting heart rate reduction on Day 3, consistent with rapid attenuation of the initial cardiac effect during repeated dosing.

Because rapid onset of bradycardia and Wenckebach-type AV conduction delays in healthy participants may foreshadow rare but clinically relevant first-dose cardiac events in larger patient populations, the present study evaluated whether two dose-titration regimens could mitigate the initial negative chronotropic effect of mocravimod while preserving its PK and PD properties.

## Material and methods

### Participants

Healthy male and female non-smokers aged 18–55 years with a body mass index of 18–30 kg/m^2^ were eligible. Health status was confirmed at screening and baseline by medical history, physical and neurological examination, vital signs (including orthostatic measurements), 12-lead electrocardiogram (ECG), clinical laboratory testing (including haematology with absolute lymphocyte count [ALC]), and pulmonary function testing by spirometry. Key exclusion criteria included clinically significant abnormalities in pulmonary function tests, ALC < 1.0 × 10^9^/L at screening and/or baseline, clinically relevant cardiovascular or ECG abnormalities (including relevant AV conduction abnormalities and QT prolongation), pregnancy or lactation, current smoking (urine cotinine ≥ 500 ng/mL), recent use of investigational drugs, and use of prohibited concomitant medications prior to first dosing.

### Study design

This was a single-centre, randomised, double-blind, placebo-controlled, parallel-group, multiple-dose phase I study in healthy participants. The study was designed to determine whether initial dose titration of mocravimod resulted in fewer bradycardia episodes on treatment initiation and dose-increase days than fixed initiation at 2 mg once daily (Figure 1). After a screening period of up to 21 days, participants entered the clinic on Day −2, received placebo on Day −1 during the run-in period, and received once-daily oral study treatment from Day 1 through Day 21. On Day −1, all participants underwent baseline assessments, including 24-hour Holter monitoring and ambulatory blood pressure monitoring (ABPM). Participants were randomized, stratified by baseline pulse rate (50–59 bpm or ≥60 bpm), to one of four groups: dose titration group 1 (DT1), dose titration group 2 (DT2), fixed-dose mocravimod, or placebo. DT1 received 0.3, 0.6, 0.9, 1.2, and 2.0 mg once daily, with dose escalation every four days. DT2 received 0.5 mg once daily through Day 14, 1.2 mg once daily from Days 15 to 18, and 2.0 mg once daily thereafter. The fixed-dose group received 2.0 mg once daily throughout the 21-day treatment period. Study drug doses and dose-escalation steps are shown in Figure 1. Study medication was administered between approximately 08:30 and 09:30 after a modest breakfast served between approximately 07:30 and 08:30. Participants remained in-house through Day 22 and returned for follow-up visits on Days 35 and 42.

**Figure 1.**
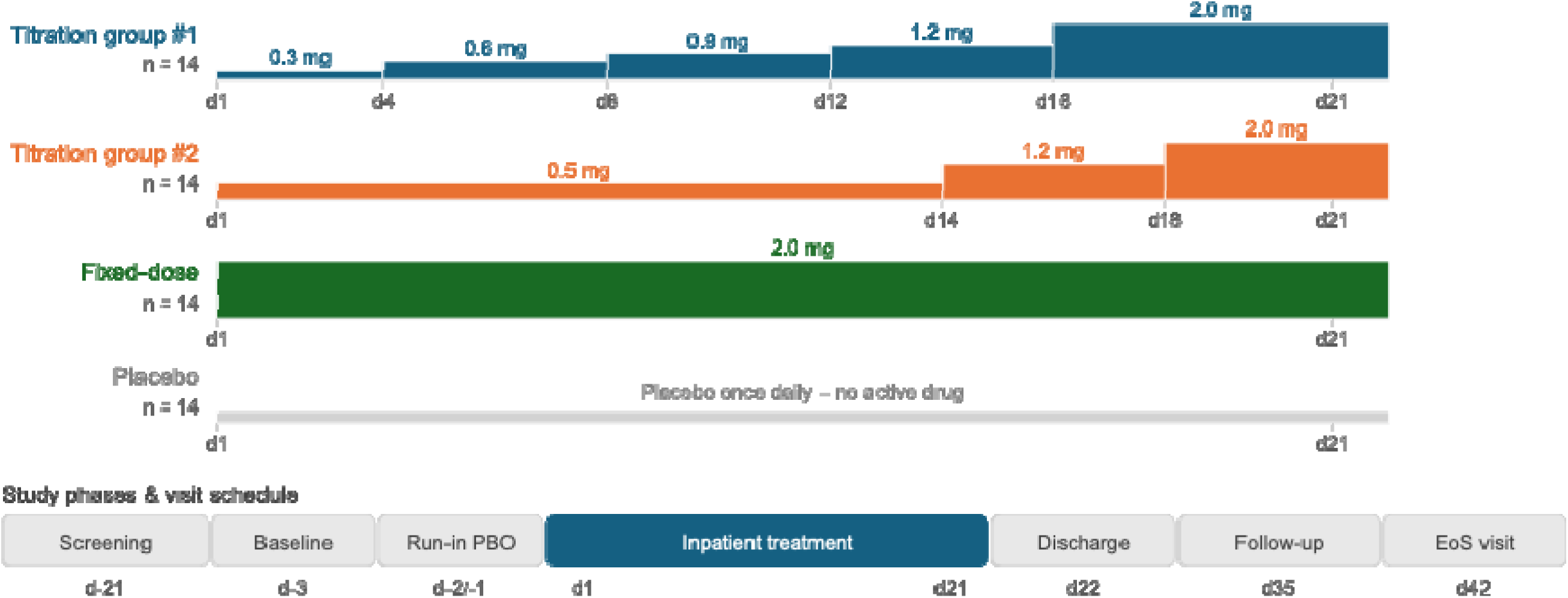
Study design. Four parallel groups (n=14 each) received once-daily study drug over a 21-day inpatient period. Each panel below shows one group’s daily dose: bars rise as the dose increases toward the 2.0 mg target. d, day; EoT, end-of-treatment; EoS, end-of-study; PBO, placebo; qd, once daily

The study protocol was reviewed by the Independent Ethics Committee (IEC) for the centre, i.e., the South East Wales Research Ethics Committee Panel B, Cardiff, United Kingdom. The study was conducted according to the ethical principles of the Declaration of Helsinki. Informed consent was obtained from each participant in writing before randomization.

### Bioanalysis

PK assessments included mocravimod and its active metabolite, mocravimod-phosphate, measured in whole blood. Blood samples were collected in EDTA tubes via venipuncture or an indwelling cannula, mixed by gentle inversion, split into aliquots, and stored frozen at −70°C or below until analysis. Serial PK profiles were collected on Day 1 at pre-dose and 0.5, 3, 6, 10, and 12 h post-dose; on Days 2, 5, 9, 13, 15, 17, and 19 at pre-dose and 3, 6, 10, and 12 h post-dose; and on Day 21 at pre-dose and 3, 6, 10, 12, and 24 h post-dose. Additional pre-dose samples were collected on Days 3, 6, 10, 14, 16, 18, and 20, with one follow-up sample on Day 36. Mocravimod and mocravimod-phosphate concentrations were quantified using a validated LC–MS/MS method with a lower limit of quantification of 0.05 ng/mL for both analytes. The method was not enantioselective; therefore, mocravimod-phosphate was measured as the racemate.

### Safety assessments

Safety was assessed throughout the study by adverse event and serious adverse event monitoring, physical examinations, vital signs (including orthostatic blood pressure on scheduled days), body weight, clinical laboratory tests (haematology, clinical chemistry, and urinalysis), pregnancy testing, and pulmonary function tests. Cardiac safety and pharmacodynamic effects were monitored using standard 12-lead ECGs and continuous 24-hour 12-lead Holter recordings, with additional 24-hour ambulatory blood pressure monitoring. Adverse events were collected via subject reporting, non-directive questioning, and investigator observation and were followed until resolution or stabilization.

### Statistical analysis

The primary endpoint was the daily number of bradycardia episodes, as assessed by 24-hour Holter monitoring. The primary comparison evaluated the number of bradycardia episodes on dose-escalation days in the titration regimens (Days 1, 5, 9, 13, and 17 for DT1; Days 1, 15, and 19 for DT2) against Day 1 in the fixed-dose 2 mg group. In addition, the total number of bradycardia episodes during the treatment period was compared between treatment groups. Pairwise comparisons included each titration regimen versus the fixed-dose regimen, DT1 versus DT2, and each active treatment group versus placebo. As the endpoint comprised count data with a non-normal distribution, comparisons were performed using a non-parametric rank-based test according to Neubert and Brunner. Mean ranks, variances, and permutation-based *P* values were calculated for each pairwise comparison, with lower mean ranks indicating fewer bradycardia episodes. A bradycardia episode was defined as a one-minute interval with heart rate <40 bpm during daytime (07:00–22:00) or <35 bpm during night-time (22:00–07:00), or heart rate <50 bpm during daytime or <45 bpm during night-time with a concomitant reduction of ≥15 bpm compared with the matching hourly average baseline heart rate. Baseline was defined as the matching hourly average heart rate from Days −2 and −1.

The prespecified analysis of the primary endpoint used a repeated Poisson model with day, treatment, and treatment-by-day interaction as fixed effects and generalized estimating equations to account for repeated observations, with adjustment for overdispersion. Because this model and an alternative negative-binomial model did not adequately fit the observed event distribution (of the data (highproportion of zero values for placebo, highly asymmetrical and heterogeneous event distribution for the active treatment groups on the different days), the primary analysis was performed using a non-parametric rank-based comparison of treatment groups with respect to the daily number of bradycardia episodes. The same approach was used to compare the total number of bradycardia episodes during the treatment period. Hourly minimum and mean heart rate and their ratios to baseline were analysed on the natural logarithmic scale using mixed-effects linear models with fixed circadian, treatment-by-day effects and participant-level random effects on the intercept and circadian effect. Estimates and 95% confidence intervals were back-transformed to the natural scale.

Secondary endpoints included bradycardia episodes during each titration or fixed-dose regimen compared with placebo; the frequency of second-degree atrioventricular blocks and sinus pauses >3 s; hourly minimum and mean heart rate and their changes from baseline; safety and tolerability; pharmacokinetics of mocravimod and mocravimod-phosphate; and absolute lymphocyte count as a pharmacodynamic marker. Event-based secondary endpoints were summarized by treatment group and study day. Pharmacokinetic parameters were derived from whole-blood concentration–time data using non-compartmental analysis, including AUClast, AUC0–24h, Cmax, Tmax, accumulation ratios, and molar mocravimod-phosphate/mocravimod exposure ratios, and were summarized descriptively. Safety outcomes were summarized descriptively by treatment group. The safety population included all participants who received at least one dose of study drug; PK and PD populations included participants with evaluable PK and PD data, respectively.

## Results

### Disposition

A total of 56 participants were enrolled and randomized into the four treatment groups, and 53 completed the study. Three participants discontinued, two due to AEs and one was lost to follow-up.

### Demographic characteristics

Study participants were healthy males with a median age of 27 (18–52) years. The majority (n=53; 94.6%) were of Caucasian origin (Table 1).

**Table 1.** Demographics by treatment group.

|  |  | **Titration #1** | **Titration #2** | **Fixed dose** | **Placebo** | **Total** |
| --- | --- | --- | --- | --- | --- | --- |
|  |  | **N=13** | **N=14** | **N=14** | **N=15** | **N=56** |
| Age (years) | Mean (SD) | 31.8 (11.3) | 29.1 (8.6) | 27.9 (5.0) | 27.9 (7.0) | 29.1 (8.1) |
|  | Median | 27.0 | 27.5 | 26.5 | 26.0 | 27.0 |
|  | Range | 19 – 52 | 18 – 45 | 22 – 38 | 18 – 42 | 18 – 52 |
| Gender – n (%) | Male | 13 (100) | 14 (100) | 14 (100) | 15 (100) | 56 (100) |
| Race – n (%) | Caucasian | 11 (84.6) | 13 (92.9) | 14 (100) | 15 (100) | 53 (94.6) |
|  | Other | 2 (15.4) | 1 (7.1 %) | – | – | 3 (5.4) |
| Ethnicity – n (%) | Indian (India subc) | 1 (7.7) | – | – | – | 1 (1.8) |
|  | Mixed ethnicity | 1 (7.7) | – | – | – | 1 (1.8) |
|  | Other | 11 (84.6) | 14 (100) | 14 (100) | 15 (100) | 54 (96.4) |
| Baseline supine pulse rate (bpm) | Mean (SD) | 63.9 (5.7) | 64.9 (8.3) | 64.5 (10.0) | 64.0 (11.3) | 64.3 (8.9) |
|  | Median | 62.0 | 65.5 | 65.0 | 62.0 | 64.0 |
|  | Range | 57 - 76 | 52 - 81 | 49 - 81 | 47 - 98 | 47 - 98 |
| Body Mass Index<br>(kg/m<sup>2</sup>) | Mean (SD) | 26.30 (2.41) | 26.13 (2.39) | 25.58 (2.11) | 24.38 (2.18) | 25.57 (2.34) |
|  | Median | 26.41 | 26.31 | 25.64 | 24.07 | 25.73 |
|  | Range | 21.7 – 29.4 | 22.8 – 29.9 | 22.3 – 28.3 | 21.2 – 27.9 | 21.2 – 29.9 |
| Weight (kg) | Mean (SD) | 80.18 (8.78) | 83.12 (6.94) | 79.50 (9.58) | 76.51 (6.06) | 79.76 (8.06) |
|  | Median | 79.50 | 84.45 | 79.70 | 75.40 | 79.65 |
|  | Range | 64.2 – 91.7 | 68.9 – 94.8 | 65.3 – 100.9 | 68.8 – 89.6 | 64.2 – 100.9 |
| Height (cm) | Mean (SD) | 174.5 (5.1) | 178.5 (5.9) | 176.1 (6.8) | 177.3 (6.4) | 176.7 (6.1) |
|  | Median | 173.0 | 177.5 | 174.5 | 179.0 | 176.5 |
|  | Range | 166 – 184 | 172 – 194 | 167 – 189 | 166 – 187 | 166 – 194 |

SD, standard deviation; subc, subcontinent

### Pharmacokinetics

PK profiles of mocravimod and mocravimod-phosphate were consistent with expected drug levels based on previous phase I single and multiple dose data [5, 6]. None of the two dose-titration regimens led to PK steady-state. Steady-state was only reached in the fixed 2 mg dose arm between Day 14 and Day 21, consistent with the results from study CKRP203A2102 [6]. By Day 21, blood concentrations in DT1 were close to those of the 2 mg fixed dose group and of similar magnitude, whereas DT2 yielded lower exposures (Figure 2, Table 2). Inter-participant variability remained low to moderate regardless of the treatment arm and was not affected by dose increments. The PK profile of mocravimod-phosphate followed the same pattern of the parent mocravimod. Molar mocravimod/mocravimod-phosphate exposure ratios remained close to 1 throughout the entire treatment period in all arms.

**Figure 2.**
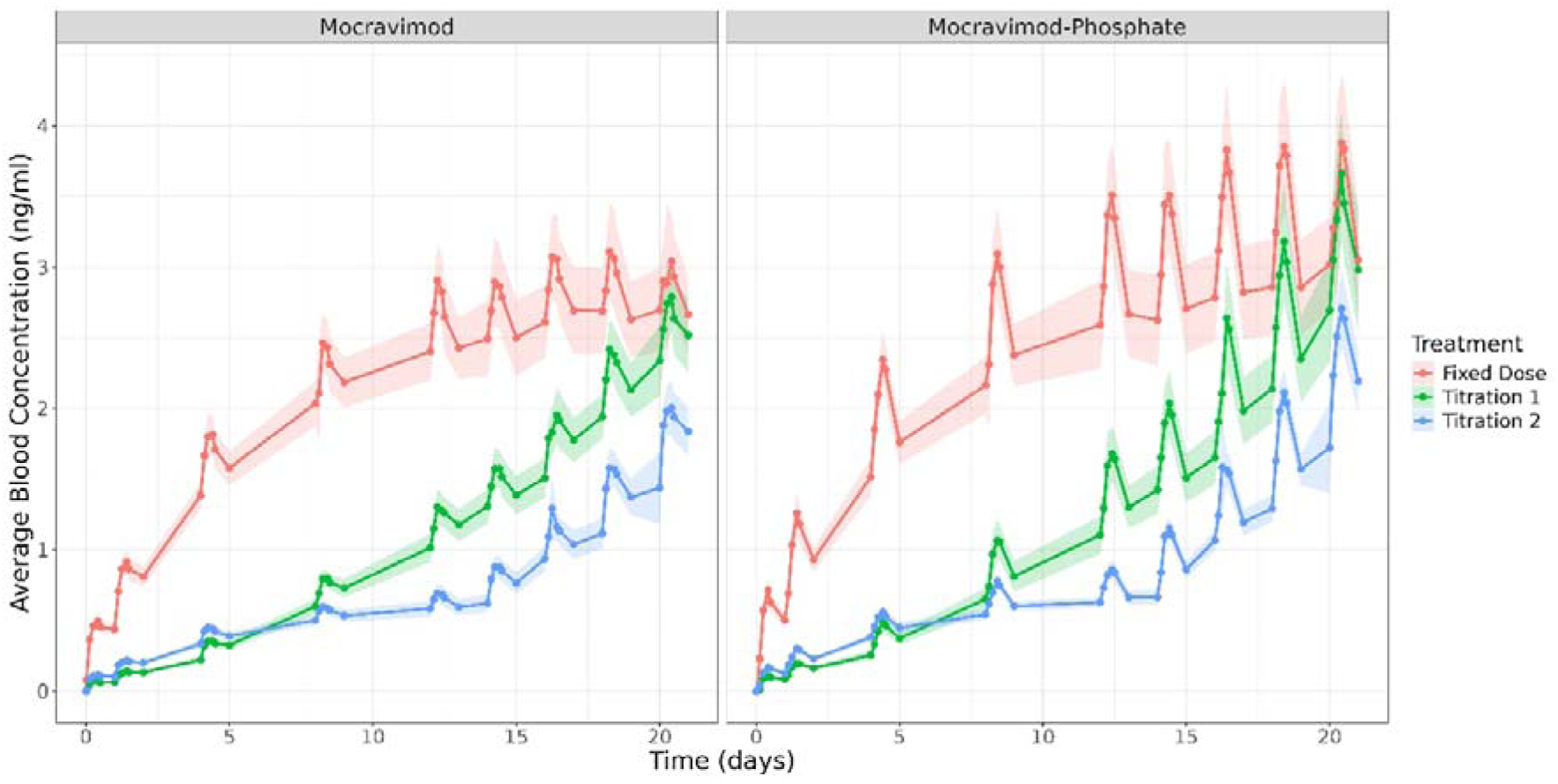
Average blood concentrations over time. Results are shown and mean ± 95% confidence intervals.

**Table 2.** PK parameters by treatment group.

| **Day** | **Analyte** | **Parameter** | **Titration #1**<br>**N=13** | **Titration #2**<br>**N=14** | **Fixed dose**<br>**N=14** |
| --- | --- | --- | --- | --- | --- |
| 1 | Mocravimod | Tmax (hr) | 10.00 (3.00 – 23.98) | 10.00 (6.00 – 23.97) | 10.00 (3.00 – 23.97) |
|  |  | Cmax (ng/ml) | 0.08 (35.7%) | 0.12 (12.4%) | 0.50 (15.4%) |
|  |  | AUC0-24 (hr.ng/mL) | 1.45 (19.7%) | 2.32 (9.4%) | 9.90 (15.5%) |
|  | Mocravimod-Phosphate | Tmax (hr) | 10.00 (6.02 – 23.97) | 10.00 (6.00 – 12.00) | 10.00 (10.00 – 12.00) |
|  |  | Cmax (ng/ml) | 0.11 (32.0%) | 0.17 (20.6%) | 0.70 (23.1%) |
|  |  | AUC0-24 (hr.ng/mL) | 1.78 (29.3%) | 2.87 (21.2%) | 11.94 (21.1%) |
| 21 | Mocravimod | Tmax (hr) | 10.00 (3.00 – 12.00) | 10.00 (3.00 – 12.00) | 6.00 (3.00 – 12.00) |
|  |  | Cmax (ng/ml) | 2.79 (16.6%) | 2.01 (17.9%) | 3.06 (19.9%) |
|  |  | AUC0-24 (hr.ng/mL) | 61.86 (17.3%) | 44.77 (17.1%) | 67.15 (20.9%) |
|  | Mocravimod-Phosphate | Tmax (hr) | 10.00 (6.00 – 12.00) | 10.00 (3.00 – 12.00) | 10.00 (6.00 – 12.00) |
|  |  | Cmax (ng/ml) | 3.64 (21.9%) | 2.74 (19.5%) | 3.90 (23.3%) |
|  |  | AUC0-24 (hr.ng/mL) | 76.08 (23.0%) | 56.95 (18.1%) | 81.31 (22.5%) |

### Number of bradycardia events

The boxplots of the number of bradycardia episodes per day by treatment group and study visit are provided in Figure 3. On Day 1, both titration regimens had less bradycardic episodes compared to the fixed dose. The difference between the two titration regimens and the fixed dose was not observed after Day 9 (Figure 3). The non-parametric comparison of the total number of bradycardia episodes during the treatment period demonstrated differences between the treatment regimens. Subjects receiving the fixed-dose regimen had significantly more bradycardia episodes than those receiving DT2 (mean rank 18.3 vs. 10.7; permutation *P* = 0.019) and placebo (mean rank 20.4 vs. 9.9; permutation *P* = 0.001) (Table 3). No significant difference was observed between DT2 and placebo (mean rank 16.3 vs. 13.8; permutation *P* = 0.407). DT1 showed intermediate results, with numerically fewer bradycardia episodes than the fixed-dose regimen (mean rank 11.8 vs. 16.0; permutation *P* = 0.174) and numerically more episodes than placebo (mean rank 17.6 vs. 11.8; permutation *P* = 0.128), although neither comparison reached statistical significance. Likewise, no significant difference was observed between the two titration regimens (mean rank 15.8 vs. 12.3; permutation *P* = 0.287). Overall, these findings suggest that the titration strategy, particularly DT2, reduced the occurrence of bradycardia episodes compared with the fixed-dose regimen, whereas the incidence observed with DT2 was comparable to placebo.

**Figure 3.**
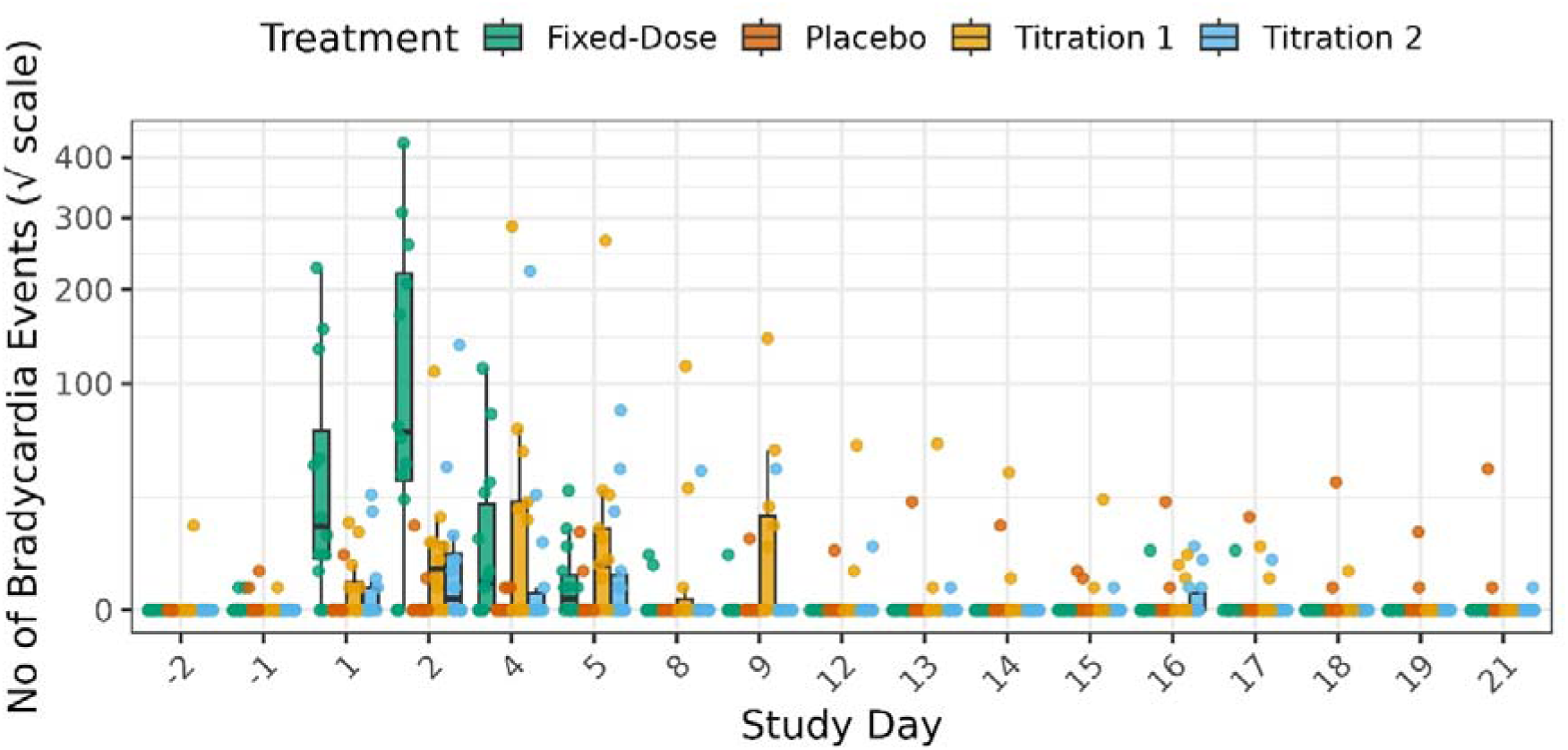
Boxplots of bradycardia episodes per treatment by study day.

**Table 3.** Non-parametric test for the comparison of the total number of bradycardia episodes during the treatment period.

| Comparison |  |  | Rank means |  | Variance |  | Pperm | Pt |
| --- | --- | --- | --- | --- | --- | --- | --- | --- |
| Treatment (1) | Treatment (2) | N (N1 vs N2) | R1 | R2 | Var1 | Var2 |  |  |
| Titration 1 | Titration 2 | 27 (13 vs 14) | 15.8 | 12.3 | 18.97 | 11.37 | 0.287 | 0.245 |
| Titration 1 | Fixed dose | 27 (13 vs 14) | 11.8 | 16.0 | 18.14 | 12.27 | 0.174 | 0.178 |
| Titration 1 | Placebo | 28 (13 vs 15) | 17.6 | 11.8 | 23.24 | 7.20 | 0.128 | 0.062 |
| Titration 2 | Fixed dose | 28 (14 vs 14) | 10.7 | 18.3 | 13.26 | 12.03 | 0.019 | 0.009 |
| Titration 2 | Placebo | 29 (14 vs 15) | 16.3 | 13.8 | 22.25 | 11.10 | 0.407 | 0.406 |
| Fixed dose | Placebo | 29 (14 vs 15) | 20.4 | 9.9 | 14.53 | 4.07 | 0.001 | 0.000 |

### Other endpoints

A total of 5 participants developed 2°AV block Type I: two participants from DT1 (one pre-dose only and the other pre- & post-dose), one participant from DT2 (pre-dose only), one participant from the fixed dose (post-dose), and one participant on placebo (post-dose). One participant in DT1 was noted with sinus pause > 3 secs on study Day 1 (max pause = 5.87 sec). This event was associated with a difficult venipuncture resulting in vasovagal episode and was considered by the investigator to be non-drug related.

The hourly minimum heart rate decreases from baseline on Day 1 and Day 2 was more evident in the fixed dose regimen compared to the two titration regimens and by Day 4 the decrease was similar in all active treatment groups. By Day 13, no decrease in heart rate compared to the matched baseline was apparent for any of the treatment groups (Figure S1).

ALC decreased from baseline to Day 21 in all the active treatment groups. On day 21, the decrease was comparable among the active treatment groups and returned to near baseline at the end of the study (Figure 4).

**Figure 4.**
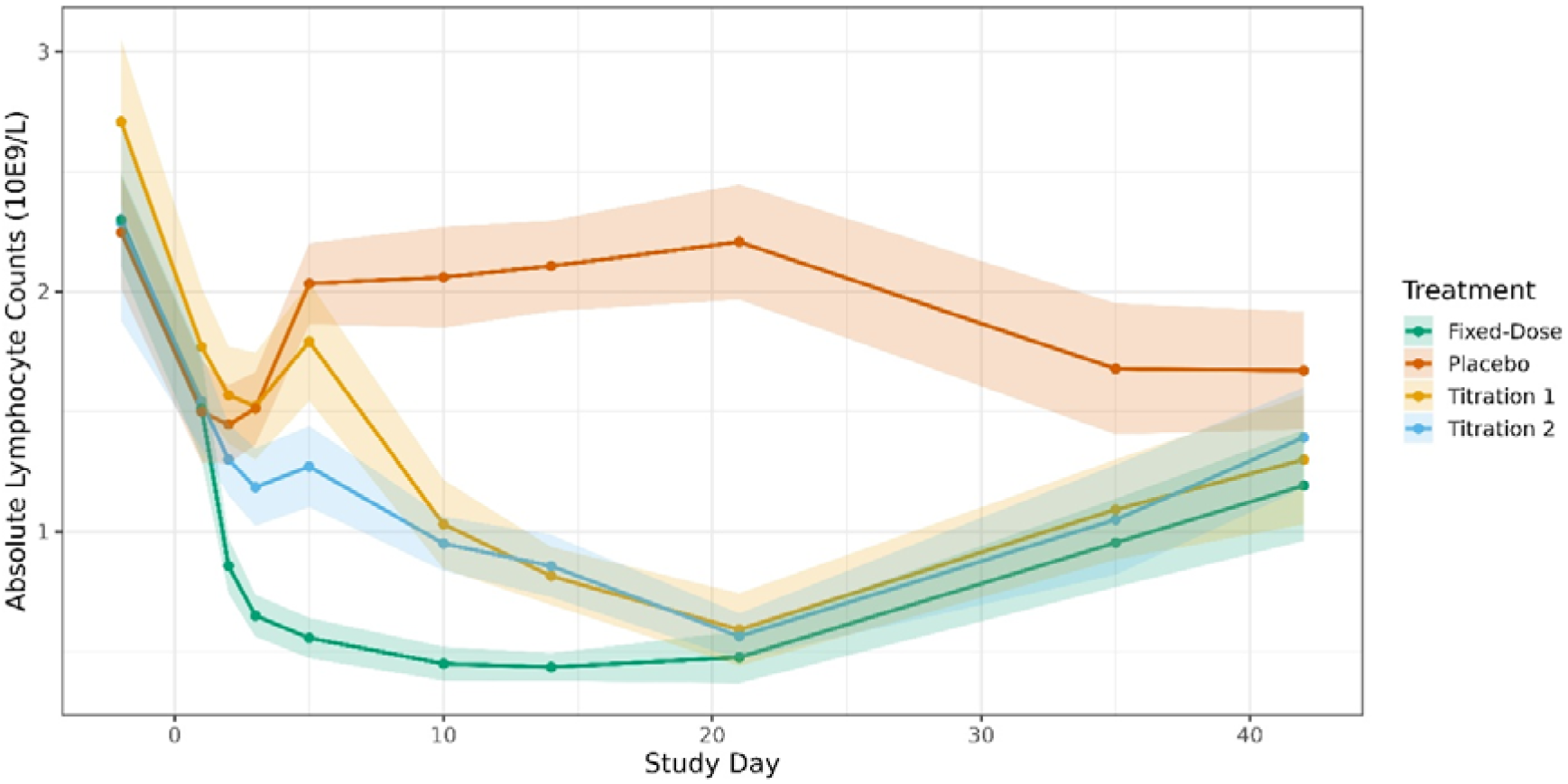
Absolute lymphocyte counts over time. Results are shown and mean ± 95% confidence intervals.

### Safety results

No serious adverse events (SAEs) were reported. A total of 48 (85.7%) participants experienced at least one AE during the study (Table 4). Most of these AEs were administrative site conditions (medical device site reactions; 42.9%), nervous system disorder (37.5%), gastrointestinal disorders (25.0%) and respiratory, thoracic/mediastinal disorders (19.6%). No discernible difference in the AE profile was observed between the mocravimod dose titration groups and the placebo group. Elevated liver enzymes without bilirubin elevation were observed in 2 participants and led to their discontinuation from the study. One participant was in the fixed dose group and the other was on placebo, who was given acetaminophen for toothaches. ABPM on Day 1, Day 9, and Day 21 did not show clinically relevant changes other than reduction in heart rate (as detected by Holter monitor). One participant in DT 1 experienced a brief syncopal episode on day 1 (6 hours post-dosing) associated with difficult venipuncture. It resolved in less than one minute and was considered by the Investigator as not suspected to be related to study drug. There were no relevant changes in spirometric values associated with administered mocravimod in any of the three treatment regimens.

**Table 4.** Incidence of AEs by primary system organ class - n (%) of subjects.

|  | **Titration #1** | **Titration #2** | **Fixed dose** | **Placebo** | **Total** |
| --- | --- | --- | --- | --- | --- |
|  | **N=13** | **N=14** | **N=14** | **N=15** | **N=56** |
| Subjects with AE(s) | 10 (76.9) | 12 (85.7) | 14 (100) | 12 (80) | 48 (85.7) |
| **System organ class** |  |  |  |  |  |
| General disorders and administration site conditions | 3 (23.1) | 5 (35.7) | 9 (64.3) | 7 (46.7) | 24 (42.9) |
| Nervous system disorders | 7 (53.8) | 2 (14.3) | 8 (57.1) | 4 (26.7) | 21 (37.5) |
| Gastrointestinal disorders | 4 (30.8) | 4 (28.6) | 1 (7.1) | 5 (33.3) | 14 (25.0) |
| Respiratory, thoracic and mediastinal disorders | 4 (30.8) | 3 (21.4) | 3 (21.4) | 1 (6.7) | 11 (19.6) |
| Infections and infestations | 2 (15.4) | 1 (7.1) | – | 2 (13.3) | 5 (8.9) |
| Musculoskeletal and connective tissue disorders | – | 1 (7.1) | – | 3 (20.0) | 4 (7.1) |
| Eye disorders | – | 1 (7.1) | 1 (7.1) | 1 (6.7) | 3 (5.4) |
| Skin and subcutaneous tissue disorders | – | 1 (7.1) | 2 (14.3) | – | 3 (5.4) |
| Investigations | – | – | 1 (7.1) | 1 (6.7) | 2 (3.6) |
| Psychiatric disorders | 2 (15.4) | – | – | – | 2 (3.6) |
| Blood and lymphatic system disorders |  |  | 1 (7.1) |  | 1 (1.8) |
| Injury, poisoning and procedural complications |  |  | 1 (7.1) |  | 1 (1.8) |
| Reproductive system and breast disorders |  | 1 (7.1) |  |  | 1 (1.8) |

## Discussion

In this randomized, double-blind study in healthy participants, both dose-titration regimens mitigated the early negative chronotropic effect of mocravimod relative to initiation at the target dose of 2 mg once daily, while preserving the expected pharmacokinetic and pharmacodynamic profile. These findings align with the established class effect of S1PR modulators, where a transient reduction in heart rate and occasional, usually self-limited, AV conduction delays can occur upon treatment initiation due to functional agonism of S1PR1 on cardiac myocytes prior to receptor internalization and desensitization [7].

Across approved S1PR modulators, first-dose bradycardia is dose-related and most prominent during the initial days of exposure, with attenuation over time as tolerance develops. For some agents, this early cardiac effect is mitigated through predefined titration schedules (e.g., ozanimod), whereas for others titration is not required (e.g., etrasimod), reflecting differences in receptor selectivity, pharmacokinetics, and functional agonism at S1PR subtypes [8]. The present results place mocravimod within this broader experience and demonstrate that gradual dose escalation can meaningfully reduce the burden of bradycardia episodes observed after starting at the full 2 mg dose.

Prior early-phase evaluations of mocravimod reported clinically relevant, transient reductions in heart rate following initiation at higher doses [5, 6], consistent with the S1PR class effect . The current study assessed whether two practical titration regimens (DT1 and DT2) could reduce the incidence of protocol-defined bradycardia episodes on dose initiation and dose-escalation days compared with fixed initiation at 2 mg once daily. Both titration regimens resulted in fewer bradycardia episodes on Day 1 than the fixed-dose regimen, and differences between titration and fixed-dose were no longer evident after Day 9, consistent with the development of tolerance with continued daily dosing.

Second-degree AV block Type I events were infrequent, occurred across active arms as well as placebo, and events observed in the titration regimens were not suspected to be study-drug related. Analyses of hourly minimum and mean heart rate indicated a more pronounced decrease early after treatment start with the fixed-dose regimen, while heart rate reductions in the titration groups were smaller initially and converged by Day 4. By Day 13, no decrease versus matched baseline was apparent in any active treatment group, supporting that the negative chronotropic effect is transient and diminishes with ongoing exposure. Beyond mitigating early bradycardia, an important consideration is whether titration preserves the intended exposure and pharmacodynamic activity. In this study, both titration regimens maintained the characteristic mocravimod and mocravimod-phosphate exposure relationship and produced a reduction in absolute lymphocyte count comparable to fixed-dose treatment by Day 21, indicating that the pharmacodynamic effect was preserved. DT1 achieved concentrations close to those observed with 2 mg fixed dose by the end of dosing, whereas DT2 resulted in lower exposures, consistent with the planned slower escalation and supporting its use as a more conservative initiation option when additional mitigation of early cardiac effects is desired. Clinically, the choice of titration schedule should balance cardiovascular tolerability against the need to achieve therapeutic exposure rapidly. For chronic indications such as multiple sclerosis or inflammatory bowel disease, a slower escalation may be acceptable and operationally straightforward. In contrast, in settings where rapid attainment of target exposure is prioritized, such as transplant immune modulation in allo-HCT, shorter titration approaches or alternative initiation strategies may be preferred [9, 10]. Mocravimod is currently being evaluated in a large, randomized placebo-controlled phase 3 study to boost graft versus leukaemia effect in patients undergoing allo-HCT for acute myeloid leukaemia (NCT05429632) [11].

## Conclusion

Both titration regimens minimized the initial bradycardic effect observed with the 2 mg fixed dose regimen. No relevant heart-related events were observed with the two titration regimens. Both titration regimens showed a reduction in ALC comparable to that observed with the fixed dose at the end of the dosing period and returned to near baseline at the end of the study. Both titration regimens allowed reaching concentrations similar to those in the 2 mg fixed dose group at the end of the dosing period. Both titration regimens were safe and well tolerated and our findings support further clinical development.

## Data Availability

All data produced in the present study are available upon reasonable request to the authors

## Acknowledgements

The authors wish to thank Dr. Margit Hemetsberger, Vienna, Austria, for editorial services, funded by Priothera SAS, Saint-Louis, France. This study was conducted by Novartis Pharma AG, Basel, Switzerland.

## References

1. McGinley MP, Cohen JA. Sphingosine 1-phosphate receptor modulators in multiple sclerosis and other conditions. Lancet. 2021;398(10306):1184–94. doi:10.1016/s0140-6736(21)00244-0

2. Dertschnig S, Gergely P, Finke J, Schanz U, Holler E, Holtick U, et al. Mocravimod, a Selective Sphingosine-1-Phosphate Receptor Modulator, in Allogeneic Hematopoietic Stem Cell Transplantation for Malignancy. Transplantation and Cellular Therapy. 2023;29(1):41.e1–.e9. doi:10.1016/j.jtct.2022.10.029

3. Amoozgar B, Bangolo A, Mohamed A, Mansour C, Elias D, Cho C, et al. JAK2 Inhibitors and Emerging Therapies in Graft-Versus-Host Disease: Current Perspectives and Future Directions. Biomedicines. 2025;13(7):1527

4. Dekker L, Sanders E, Lindemans CA, de Koning C, Nierkens S. Naive T Cells in Graft Versus Host Disease and Graft Versus Leukemia: Innocent or Guilty? Front Immunol. 2022;13:893545. doi:10.3389/fimmu.2022.893545

5. Huntjens D, Klingbiel D, Hasskarl J. A Randomized, Double-Blind, Placebo-Controlled, Single Ascending Oral Dose Study of Mocravimod: Safety, Tolerability, Pharmacokinetics, and Pharmacodynamics in Healthy Participants. medRxiv. 2026:2026.05.11.26352861. doi:10.64898/2026.05.11.26352861

6. Huntjens D, Klingbiel D, Hasskarl J. Safety, Tolerability, Pharmacokinetics, and Pharmacodynamics of Multiple Ascending Doses of mocravimod in Healthy Volunteers. medRxiv. 2026:2026.05.22.26353846. doi:10.64898/2026.05.22.26353846

7. Constantinescu V, Haase R, Akgün K, Ziemssen T. S1P receptor modulators and the cardiovascular autonomic nervous system in multiple sclerosis: a narrative review. Ther Adv Neurol Disord. 2022;15:17562864221133163. doi:10.1177/17562864221133163

8. Choon XY, Yeo JH, White C, Sharma E, Samaan MA. The Current Sphingosine 1 Phosphate Receptor Modulators in the Management of Ulcerative Colitis. Journal of Clinical Medicine. 2025;14(10):3475

9. de Lima M, Malard F, Olavarria E, Pérez López E, Medinger M, Kueenburg E, et al. MO-TRANS: A Phase 2b Placebo-Controlled Study of Mocravimod As Adjunctive and Maintenance Treatment in AML Patients Undergoing Allogeneic Hematopoietic Cell Transplant. Blood. 2022;140(Supplement 1):12936–7. doi:10.1182/blood-2022-159785

10. Dertschnig S, Passweg J, Bucher C, Medinger M, Tzankov A. Mocravimod, a S1P receptor modulator, increases T cell counts in bone marrow biopsies from patients undergoing allogeneic hematopoietic stem cell transplantation. Cellular Immunology. 2023;388–389:104719. doi:10.1016/j.cellimm.2023.104719

11. ClinicalTrials.gov. Mocravimod as Adjunctive and Maintenance Treatment in AML Patients Undergoing Allo-HCT (MO-TRANS) 2025. Available from: https://www.clinicaltrials.gov/study/NCT05429632.

