## Supplementary figures and images for "Titration regimens mitigate mocravimod-induced negative chronotropic effect while preserving the pharmacokinetic and pharmacodynamic properties"

### Supplemental figures and tables

#### Supplemental data

##### Figure S1. Average heart rate over time per treatment group.

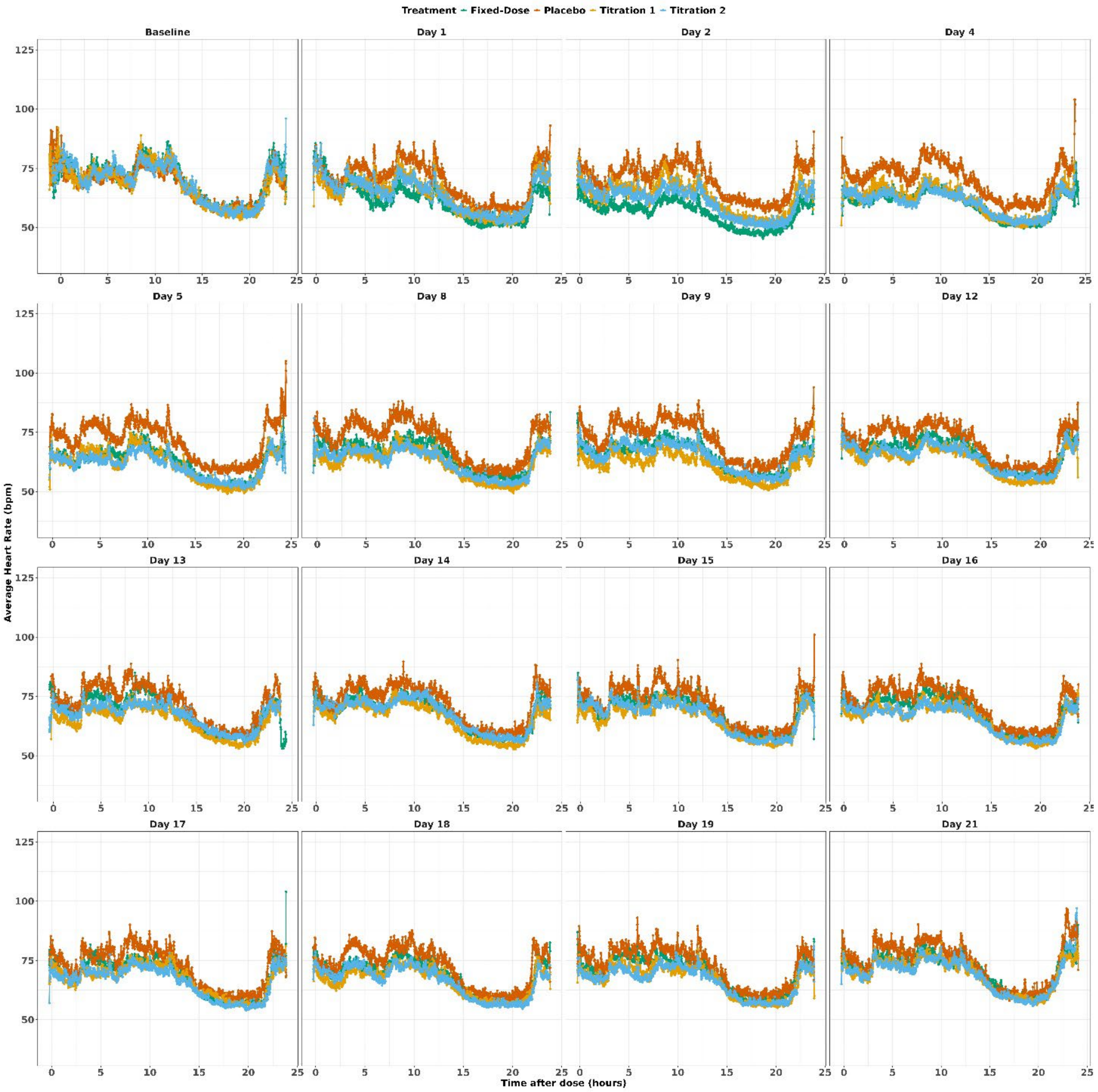
